# Development of a Soluble KIT (sKIT) Electrochemical Aptasensor For Cancer Theranostics

**DOI:** 10.1101/2020.12.04.20244186

**Authors:** Saeromi Chung, Jason K. Sicklick, Partha Ray, Drew A. Hall

**Affiliations:** Department of Electrical and Computer Engineering, University of California – San Diego, La Jolla, CA 92093, USA; Department of Surgery, Division of Surgical Oncology, Moores Cancer Center, University of California – San Diego Health, San Diego, CA 92093, USA; Department of Bioengineering, University of California – San Diego, La Jolla, CA 92093, USA

**Keywords:** KIT, aptamer, electrochemical sensor, cancer, square wave voltammetry, aptasensor, theranostics

## Abstract

A conformational changing aptamer based electrochemical sensor was developed for the detection of soluble KIT, a cancer biomarker, in human serum. The sensor was fabricated with a ferrocene labeled aptamer (*K*_d_ < 5 nM) conjugated to a gold electrode. Quantitative KIT detection was performed using electrochemical impedance spectroscopy (EIS) and square wave voltammetry (SWV). The experimental parameters such as the ratio of aptamer to spacer, aptamer immobilization time, pH, and KIT incubation time were optimized by EIS, and the sensing surface was characterized with voltammetry. The assay specificity was investigated using interfering species showing high specificity towards the target protein. The aptasensor exhibited a wide dynamic range from 10 pg/mL to 100 ng/mL in buffer with a detection limit of 1.15 pg/mL. The sensor also exhibited a linear response with increased KIT concentrations spiked in human serum. The applicability of the sensor was successfully demonstrated using cancer cell conditioned media. The proposed aptasensor can be used in continuous or intermittent approach for cancer therapy monitoring and diagnostics (theranostics).

## 1. Introduction

Cancer is a noncommunicable disease (NCD) that is responsible for most global deaths. Worldwide, one out of six deaths can be attributed to cancer – more than the mortality caused by tuberculosis, HIV/AIDS, and malaria combined. In 2018, there were ∼17 million new cases of cancer diagnosed around the world causing 9.5 million deaths (Bray et al., 2018). In the 21^st^-century, cancer is the single most significant barrier towards increasing life expectancy. Critically, many cancers are diagnosed at an advanced stage when treatments are limited, resulting in a persistent high mortality rate. Notably, several cancer serum biomarkers have been identified, such as CA-125 for ovarian cancer (Scholler and Urban, 2007), CA 19.9 for pancreatic cancer (Poruk et al., 2013), S100 for Melanoma (Harpio and Einarsson, 2004), carcinoembryonic antigen (CEA) for colorectal cancer (Nicholson et al., 2015), alpha-fetoprotein (AFP) for hepatocellular carcinoma (Arrieta et al., 2007), and prostate specific antigen (PSA) for prostate cancer (Reverri et al., 2018). The detection and treatment monitoring of cancer through biomarkers that can be performed at the point-of-care (POC), especially in low resource settings could significantly reduce cancer-related morbidity and mortality and increase the overall survival rate of patients via early and precise diagnosis (Arya and Estrela, 2018; Goossens et al., 2015, 2015; Hristova and Chan, 2019).

In this regard, the prognostic significance of the presence of KIT protein in cancer patients has received strong attention (Akin et al., 2000; DePrimo et al., 2009; Hristova and Chan, 2019; Sarlomo-Rikala et al., 1998). c-KIT is a receptor tyrosine kinase that upon binding to its ligand, Stem Cell Factor (SCF), dimerizes and initiates a cascade of signal transduction that regulates cell growth and proliferation. c-KIT is also a known proto-oncogene. Overexpression or mutations in functionally important domains of the gene are associated with cancers, including Gastrointestinal Stromal Tumor (GIST) (Akin et al., 2000; DePrimo et al., 2009; Sarlomo-Rikala et al., 1998), Mast cell disease (Systemic Mastocytosis), Acute Myeloid Leukemia (AML), Testicular Seminoma, and Melanoma (Miettinen and Lasota, 2005). In health and disease, the ligand-binding extracellular portion of the c-KIT protein is cleaved from the cell surface by the metalloprotease, tumor necrosis factor α-converting enzyme (TACE or ADAM-17) and generates a soluble KIT (sKIT) fragment (Black, 2002; Wypych et al., 1995). The resulting product, sKIT, can be detected in the serum or plasma of patients with various pathological conditions and functions as a biomarker of cancer (DePrimo et al., 2009; Kawakita et al., 1995; Makowska et al., 2009; Tajima et al., 1998; Zhong et al., 2010).

Full-length c-KIT and sKIT have been studied as a biomarker with various detection methods including flow cytometry (Banerjee et al., 2020; Naeem et al., 2002), immunohistochemical analysis (Pilloni et al., 2011), and enzyme-linked immunosorbent assay (ELISA) (Tajima et al., 1998). Although these methods provide quantitative information, they suffer disadvantages in terms of requiring sophisticated pretreatment steps, high-cost, large sample volume, expensive laboratory reagents, and sophisticated instruments for the detection process, which is not ideal as a point-of-care system (Lu et al., 2015; Jo et al., 2016). On the other hand, electrochemical detection techniques provide rapid, robust, simple, and sensitive monitoring of the interaction between a target protein and sensor surface modified with its receptor or ligand molecule (Aronoff-Spencer et al., 2016; Moon et al., 2017; Chung et al., 2018; Sun and Hall, 2019). Thus, electrochemical biosensors can be used not only to diagnose and monitor cancers, but also to provide a prognostic approach to treatment. However, there are no reports of an electrochemical biosensor for detection of sKIT to date. Thus, it is necessary to develop a KIT monitoring device with electrochemical approaches in biofluids.

Recently, electrochemical aptamer-based sensors, “aptasensors,” have drawn significant interest (Hianik and Wang, 2009; Lai et al., 2007; Lubin and Plaxco, 2010; Meirinho et al., 2016). Aptamers are single-stranded DNA or RNA oligonucleotides that are selected by an *in vitro* iterative process known as SELEX (Systematic Evolution of Ligands by Exponential Enrichment) (Ellington and Szostak, 1990; Tuerk and Gold, 1990). The nucleotides in the single-stranded aptamer molecules undergo intramolecular Watson-Crick base pairing to initially form secondary and subsequently tertiary structures to assume three-dimensional shapes that function as ligands that bind to their cognate proteins with high binding affinity and specificity, which is analogous to monoclonal antibodies (Feagin et al., 2018; Oh et al., 2010; Ray et al., 2013). This property of changing conformation of the aptamer structure upon protein binding has been utilized in electronic aptamer-based sensing devices (Tong et al., 2011). To achieve this, a redox-active molecule is covalently conjugated to the aptamer and fabricated on an electrode surface. The binding of the cognate protein to the aptamer changes its conformation, and the distance of the conjugated redox probe from the electrode (Plaxco and Soh, 2011). This in turn alters the electron transfer profile between the redox labeled aptamer and the electrode. Thus, the aptamer-protein binding event is transduced into an electronic signal that can quantitate the amount of protein present in a sample (Song et al., 2008; Willner and Zayats, 2007).

In this work, we used a DNA aptamer that was selected to bind c-KIT with high affinity and specificity (*K*_d_ < 5 nM) (Banerjee et al., 2020; Zhao et al., 2015). The aptamer was covalently conjugated to the redox-label, ferrocene (Fc), and fabricated on a gold electrode to design a novel electrochemical aptasensor as shown in **Fig. 1**. The aptasensor detected soluble KIT present in the conditioned media of cancer cell lines. This target-induced conformational change of the aptamer-based sensor has the potential to be used as a diagnostic tool to evaluate the biomarker present in the biological fluids. Such an assay, when developed, could be further coupled with injectable sensors or wearable sensors for continuous monitoring of KIT levels in response to therapy, thus closing the personalized medicine cancer treatment loop. Thus, the aptasensor can function as both a diagnostic and therapeutic response tool hence the name “theranostics.”

**Figure 1.**
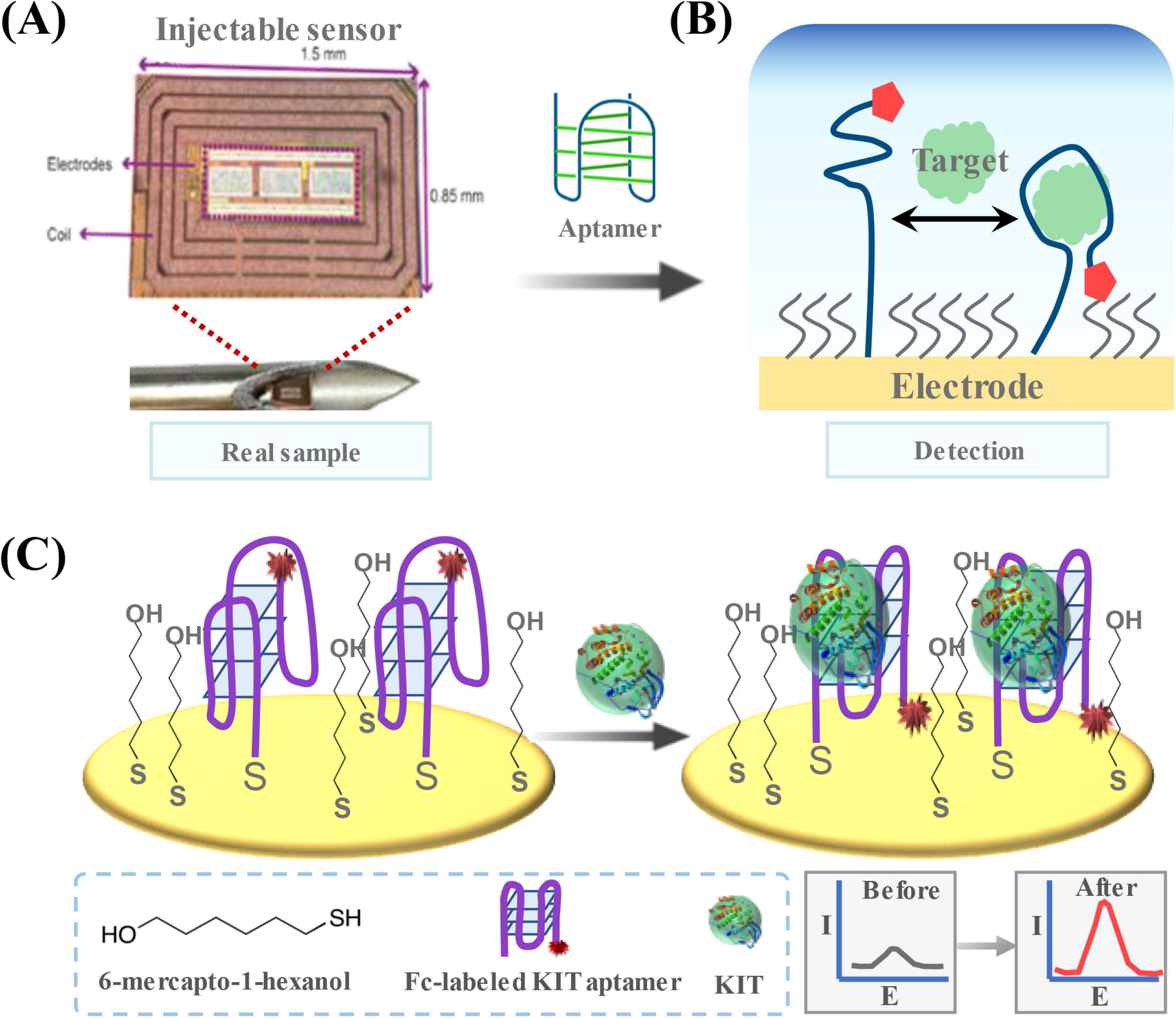
**A**. Vision of injectable sensor concept for continuous, *in vivo* cancer treatment monitoring. **B**. Conformation switching aptasensor concept. **C**. KIT aptamer with a ferrocene (Fc) label in its unbound, unfolded state (left) and bound, folded state that modulates the position of the Fc tag.

## Materials and Methods

### 2.1. Reagents and instruments

Magnesium chloride (MgCl_2_; #208337), phosphate buffered saline (PBS; #P5493), Tris[2-carboxyethyl] phosphine (TCEP; #C4706), 6-mercapto-1-hexanol (#725226), β-mercaptoethanol (#M6250), N-hydroxysuccinimide (NHS; #130672), ferrocene carboxylic acid (FCA; #106887), human serum albumin (HSA; #A9511-100MG), fibrinogen (#341576-100MG), 1,4-Dithiothreitol (DTT; #233156), Dulbecco’s Modified Eagles Medium (DMEM; #D6429-500ML), fetal bovine serum (FBS; F2442-500ML), Iscove’s Modified Dulbecco’s Medium (IMDM; 51471C-1000ML), radioimmunoprecipitation assay buffer (RIPA; R0278-50ML), and 1-thioglycerol (#M6145-25ML) were purchased from Sigma-Aldrich. Human AB serum (#01024) was bought from Omega Scientific and Ulex Europaeus Agglutinin I (UEA; #L-1060) was purchased from Vector Laboratories. Fetal Bovine serum (FBS; # A31604), Penicillin/Streptomycin (#15140122), and Pierce Protease and Phosphatase Inhibitor (#A32959) were purchased from Thermo Fisher Scientific. Interleukin 6 (IL-6; #206-IL) was purchased from R&D Systems and 3-ethylcarbodiimide (EDC; #BC25-5) from G-Biosciences. Purified c-KIT protein containing the extracellular domain (Met 1-Thr 516) with a poly-histidine tag at the C-terminus (#11996-H08H) was purchased from Sino Biological. The primary, mouse monoclonal anti-KIT antibody (#3308) was obtained from Cell Signaling Technology and the secondary, horseradish peroxidase-conjugated anti-mouse IgG (#31430) from Invitrogen. Immun-Blot PVDF membrane (#1620177) and pre-stained molecular weight marker (#1610375) were acquired from Bio-Rad.

The single-stranded DNA KIT aptamer (5’-GAG GCA TAC CAG CTT ATT CAA GGG GCC GGG GCA AGG GGG GGG TAC CGT GGT AGG ACA TAG TAA GTG CAA TCT GCG AA-3’) and the scrambled control oligo (5’-TGA CGG GAG ACT TAA AAC GCA AGG GGT GCA GCT ATC GCG GAG GCC AAG GGT TCA AGT CGA CGG GTA GCT AGG TTG GA-3’) were synthesized with 5’-S-S-(disulfide bond) and 3’-NH_2_ (amine group) modifications by Integrated DNA Technologies. We obtained the GIST-T1 cell line from T. Taguchi (Kochi Medical School, Japan), the human mast cell line HMC 1.2 from I. Pass (Sanford Burnham Prebys Medical Discovery Institute, San Diego), and the Pancreatic cancer cell line MiaPaCa-2 from ATCC (Banerjee et al., 2020).

Square Wave voltammetry (SWV), cyclic voltammetry (CV), and electrochemical impedance spectroscopy (EIS) measurements were performed using a benchtop potentiostat (CH Instruments, 750E). A 3-electrode setup was used for all experiments with Au, Ag/AgCl, and Pt wire (CH Instruments, #CHI115) as the working (WE), reference (RE), and counter electrodes (CE), respectively.

### 2.2. KIT aptasensor fabrication

Prior to electrode modification, the WE was chemically treated with a mixture of H_2_SO_4_ and H_2_O_2_ followed by washing with distilled water. It was then electrochemically cleaned by potential cycling from 0 to 1.4 V versus Ag/AgCl in 0.1 M H_2_SO_4_. The thiolated aptamer was activated with TCEP for 2 hours at room temperature. The WE was functionalized by immersing it in a solution containing 0.5 µM of aptamer in 500 µL of incubation buffer (5 mM MgCl_2_, 0.5 mM DTT in 20 mM PBS (pH 7.4)) with 0.5 µM of 6-mercapto-1-hexanol on a shaking incubator (Thermo Scientific, #4625) at room temperature for 12 hours. The WE was then gently rinsed with distilled water and equilibrated with the incubation buffer for 30 min on a shaking incubator. To remove unbound aptamer and block the surface, the electrode was incubated in 10 mM of β-mercaptoethanol for 30 min at room temperature. Lastly, the amine terminated aptamers were modified with FCA by EDC/NHS coupling (Ferapontova et al., 2008). The stepwise assembly was monitored using CV and EIS with 5 mM [Fe(CN)_6_]^4-/3-^ in 0.1 M PBS and SWV in 0.1 M PBS.

### 2.3. Circular dichroism spectroscopy

Circular dichroism (CD) experiments were performed with a CD Spectrometer (Aviv Instruments, model 202) and a quartz cuvette with a 1 mm optical path length. 400 µL of KIT aptamer (10 µM) in 100 mM potassium phosphate buffer (pH 7.1) were loaded in the cuvette and spectra was recorded from wavelengths of 220 to 320 nm in 1 nm steps at 25 °C. All scans were done in triplicate and the average of the buffer CD (baseline) was subtracted from all other measurements.

### 2.4. KIT assays

Calibration curves were generated by incubating the KIT aptasensor with various concentrations of KIT (10 pg/mL to 100 ng/mL) in 0.1 M PBS for 30 min at room temperature and measuring the response with SWV and EIS. Control assays to look at non-specific binding were performed with the control (scrambled) oligo modified sensor incubated with 10 pg/mL of KIT. Selectively was assessed using 1.0 µg/mL of IL-6, UEA I, BSA, fibrinogen, or HSA incubated for 30 min at room temperature in 0.1 M PBS (pH 7.4) and then measured using SWV and EIS. Voltammograms were recorded using SWV (−150 to +450 mV at 50 mV/s) in 0.1 M PBS (pH 7.4). EIS measurements (10 Hz to 100 kHz with a 5 mV ac amplitude) were performed with 5 mM [Fe(CN)_6_]^4-/3-^ in 0.1 M PBS.

### 2.5. Cell culturing

GIST-T1 cells were grown in DMEM with 10% FBS, 1% penicillin/streptomycin, and 2 mM glutamine (Taguchi et al., 2002). The human mast cell line HMC-1.2 was cultured in IMDM with 10% FBS, 1% penicillin/streptomycin, and 1.2 mM 1-Thioglycerol (Banerjee et al., 2020). GIST-T1 is an adherent cell line and HMC 1.2 cells grow in suspension. The growth media was collected and centrifuged at 1,000 rpm and 4 °C to remove suspended cells (HMC-1.2) or non-adherent dead cells (GIST-T1). The resulting supernatant was collected and centrifuged at 13,000 rpm and 4 °C to remove residual cellular debris. The supernatant (conditioned media, CM) was collected and stored in aliquots at -80 °C. The CM was used to estimate the soluble sKIT using an ELISA (Human CD117/c-kit Immunoassay; R&D Systems, Inc. #DSCR00).

### 2.6. Cell culture assays

For western blot, GIST-T1 cells were homogenized in RIPA buffer. 30 µL of the conditioned media and 5 µL of the GIST-cell lysate were loaded and separated by sodium dodecyl sulfate-polyacrylamide gel electrophoresis (SDS–PAGE) before transfer to a nitrocellulose membrane. Membranes were incubated with primary anti-KIT antibody (1:1,000). Secondary antibody, horseradish peroxidase-conjugated anti-mouse IgG (1:5,000) was added and antibody complexes were detected by an enhanced chemiluminescence (ECL) system (Thermo Fisher Scientific, #32109). A pre-stained molecular weight marker was run in parallel to determine the molecular weight of the proteins. For electrochemical detection, soluble KIT (sKIT) containing supernatant, which was obtained by centrifuging growth medium, was diluted to the appropriate concentration, incubated with the aptasensor for 30 min, and recorded using SWV.

## 3. Results and discussion

Guanine (G)-nucleotide rich DNA and RNA aptamers can form a four-stranded helical structure called a G-quadruplex. Four guanines connect to form a square planar structure called guanine tetrad through cyclic Hoogsteen hydrogen bonding. Two or more of these guanine tetrad layers stack to form a G-quadruplex structure. These complex structural motifs are thermodynamically stable and resistant to many serum nucleases that make them desirable candidates as therapeutic and diagnostic agents for cancer treatment (Collie and Parkinson, 2011; Tucker et al., 2012). The G-quadruplex aptamers also undergo structural change in conformation upon ligand binding and are especially suited for electrochemical aptasensor fabrication (Radi and O’Sullivan, 2006; Xiao et al., 2005).

The conformation changing aptasensor concept is illustrated in **Fig. 1B**. The sensor is functionalized with a mixture of aptamer and spacer molecules that bind to the gold electrode surface via a self-assembled chemisorption reaction between sulfur and Au forming Au-S bonds (Jo et al., 2016). In the absence of the target molecule (KIT protein in this work), the aptamer tagged with a ferrocene (Fc) reporter is in a semi-open conformation where the reporter is away from the sensor surface, as shown in **Fig. 1C**. Upon binding to KIT, the aptamer switches to a rigid, target specific G-quadruplex structure that brings the reporter near the electrode surface. Modulating the reporter position results in a decrease in electron transfer between Fc and the electrode that is readout using voltammetry techniques.

### 3.1. Aptasensor surface characterization

Each step in the aptasensor fabrication was characterized by EIS and CV starting with the unlabeled KIT aptamer (Au/aptamer). EIS was used to measure the charge transfer resistance, *R*_ct_, whereas CV measured the peak current. Spectra for a bare Au electrode, Au/aptamer, and Au/aptamer/KIT with various concentrations of KIT ranging 10 pg/mL to 10 ng/mL are shown in **Fig. 2A**. The formation of the aptamer-protein complex disrupted the electron-transfer between the ferri/ferrocyanide solution ([Fe(CN)_6_]^4-/3-^) and the sensor surface leading to an increase in *R*_ct_. The measured Nyquist plots show an *R*_ct_ increase after aptamer immobilization (Au/aptamer) from 5.07 to 8.91 kΩ. After incubating with KIT protein, the change in *R*_ct_ (Δ*R*_ct_ = *R*_ct_(KIT) - *R*_ct_(blank)) linearly increased for KIT concentrations from 10 pg/mL to 10 ng/mL. **Fig. 2B** shows the linear relationship between KIT concentration and Δ*R*_ct_ clearly demonstrating that the KIT protein interacted with the aptamer. Control experiments using the KIT aptamer (Au/aptamer) and a scrambled version of the aptamer (Au/control) further proved this. The control sensor did not show a significant Δ*R*_ct_ (0.43 ± 0.25 kΩ) thus indicating no binding compared to the KIT sensor (5.74 ± 0.36 kΩ), which had significant binding, as shown in **Fig. 2C**.

**Figure 2.**
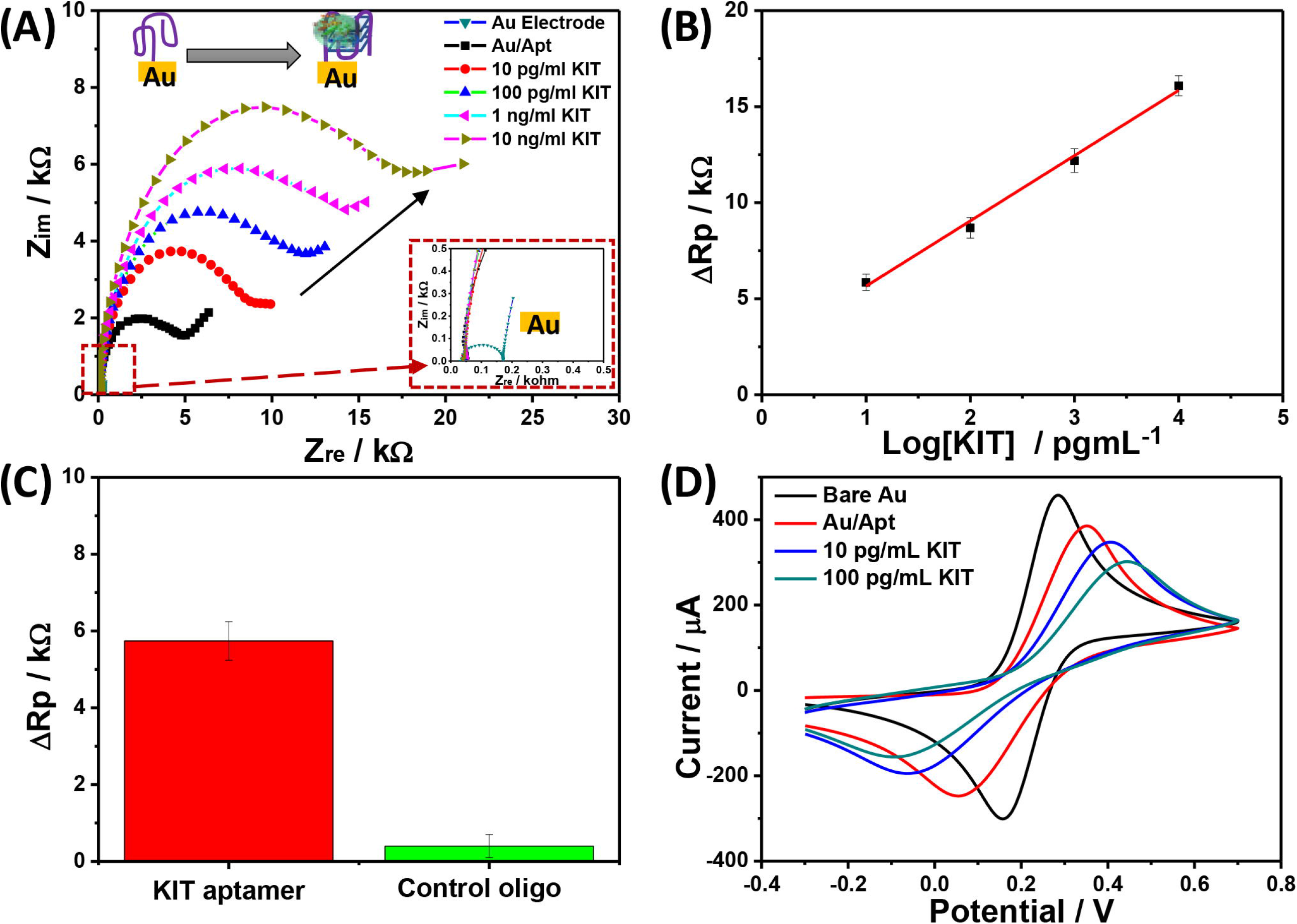
Stepwise electrode assembly. **A**. Nyquist plots of KIT aptasensor fabrication steps in 5 mM [Fe(CN)6]^4-/3-^. **B**. Extracted Δ*R*_ct_ vs. log [KIT] with a linear fit. **C**. Control experiment using KIT aptamer and scrambled oligonucleotide modified electrode with 10 pg/mL of KIT protein. **D**. Voltammograms of aptasensor fabrication in 5 mM [Fe(CN)_6_]^4-/3-^. All measurements were performed in triplicate where error bars indicate ±1σ.

CVs were also measured to assess the sensor performance. As shown in **Fig. 2D**, the voltammograms have well-defined redox peaks for [Fe(CN)_6_]^4-/3-^. The aptamer modified sensor (Au/aptamer) showed a lower peak current due to the aptamer immobilization resulting in an increase in *R*_ct_. After incubation with 10 and 100 pg/mL of KIT, the current responses decreased further because the electron transfer process between the solution and the electrode was blocked by the KIT-aptamer complex. Together, these data demonstrate that the KIT aptamer is selective towards the target molecule and that the sensor is well assembled.

To characterize the secondary aptamer structure, CD analysis was investigated in 0.1 M PBS. As shown in **Supplemental Fig. 1A**, the CD spectra has a negative peak at 245 nm and a positive peak at 270 nm, which is the inherent DNA characteristic of the G-quadruplex structure due to the multiple π-π* transitions (Kypr et al., 2009). Chronoamperometry was used to confirm the difference in electron transfer rate between the unbound and KIT bound aptamer (**Supplemental Fig. 1B**). We observed shorter exponential faradaic current decay time with KIT compared to without the KIT protein. These results demonstrate that the aptamer conformational change affects the electron transfer kinetics.

### 3.2. Aptasensor optimization

After demonstrating concept viability, we optimized the parameters (*i*.*e*., aptamer to spacer ratio, aptamer immobilization time, pH, and KIT incubation time) to obtain the best sensor response. It is well known that the ratio between the aptamer and spacer (6-mercapto-1-hexanol) molecules in conformation changing assays affects the performance as the spacer provides room to form the aptamer-protein complex. Aptamer to spacer ratios from 1:0 to 1:50 were investigated at a fixed KIT concentration (10 ng/mL). The measured Δ*R*_ct_ monotonically increased from a 1:0 to a 1:1 ratio and then declined (**Supplemental Fig. 2A**). From these data, the 1:1 ratio was chosen for all subsequent experiments. Next the aptamer incubation time was studied by incubating 0.5 µM of aptamer on the electrode from 1 to 24 hours. Δ*R*_ct_ consistently increased from 1 to 12 hours and then plateaued indicating that the sensor surface was saturated by the aptamer and spacer molecules (**Supplemental Fig. 2B**). The pH effect was examined over the range of 6.0 ∼ 8.0 (**Supplemental Fig. 2C**). The response increased gradually from 6.0 to 7.4, whereas it rapidly decreased at higher pH, likely due to the protein stability. The optimum pH for all subsequent experiments was selected to be 7.4, similar to physiological biofluid levels. Lastly, the effect of target incubation time was studied ranging from 5 to 40 min, as shown in **Supplemental Fig. 2D**. It is worth mentioning that in our envisioned *in vivo* application for treatment monitoring, this does not apply, but we did so for completeness. In these experiments, Δ*R*_ct_ monotonically increased up to 30 min incubation and then plateaued. From these results, the optimized conditions were a 1:1 ratio of aptamer to spacer, 12-hour aptamer immobilization, an assay pH of 7.4, and a 30 min sample incubation time. These values were used for all further experiments.

### 3.3. Ferrocene reporter

The assay described so far has used a traditional route with a redox couple in the buffer, ferri-/ferro-cyanide in this case. To remove the need for the redox couple towards “reagent-less”, *in vivo* monitoring, we modified the amine terminus of the aptamer with a redox reporter, ferrocene (Fc). The electrochemical behavior of each step in the fabrication process was investigated using SWV in PBS with the optimized parameters previously described, as shown in **Fig. 3**. Voltammograms were recorded for both the KIT aptamer and a scrambled version as a negative control. The ferrocene oxidation peak is clearly visible at 123 mV for the Au/Fc-Apt due to the electrochemical signal of Fc, whereas it was not observed for the bare Au, Au/spacer, or Au/aptamer modification steps. As expected, after incubating with 100 pg/mL of KIT, the Au/Fc-Apt modified electrode had an increased peak current (0.668 µA) compared to without KIT (0.262 µA) due to the conformation change of the aptamer modulating the location of the Fc reporter (**Fig. 3A**). The scrambled control oligonucleotide was tested in the same manner. As shown in **Fig. 3B**, the voltammograms without the Fc reporter (*i*.*e*. Au/control and Au/control/KIT) did not exhibit an appreciable current response. After labeling the control oligo with the Fc reporter (Au/Fc-control), the Fc oxidation peak at 128 mV was present but did not increase after incubation with 100 pg/mL of KIT, indicating that the control oligo is not binding to KIT. These results provide further evidence that the aptamer conformation change is specific to KIT and that the aptasensor can be readout using the Fc reporter and SWV.

**Figure 3.**
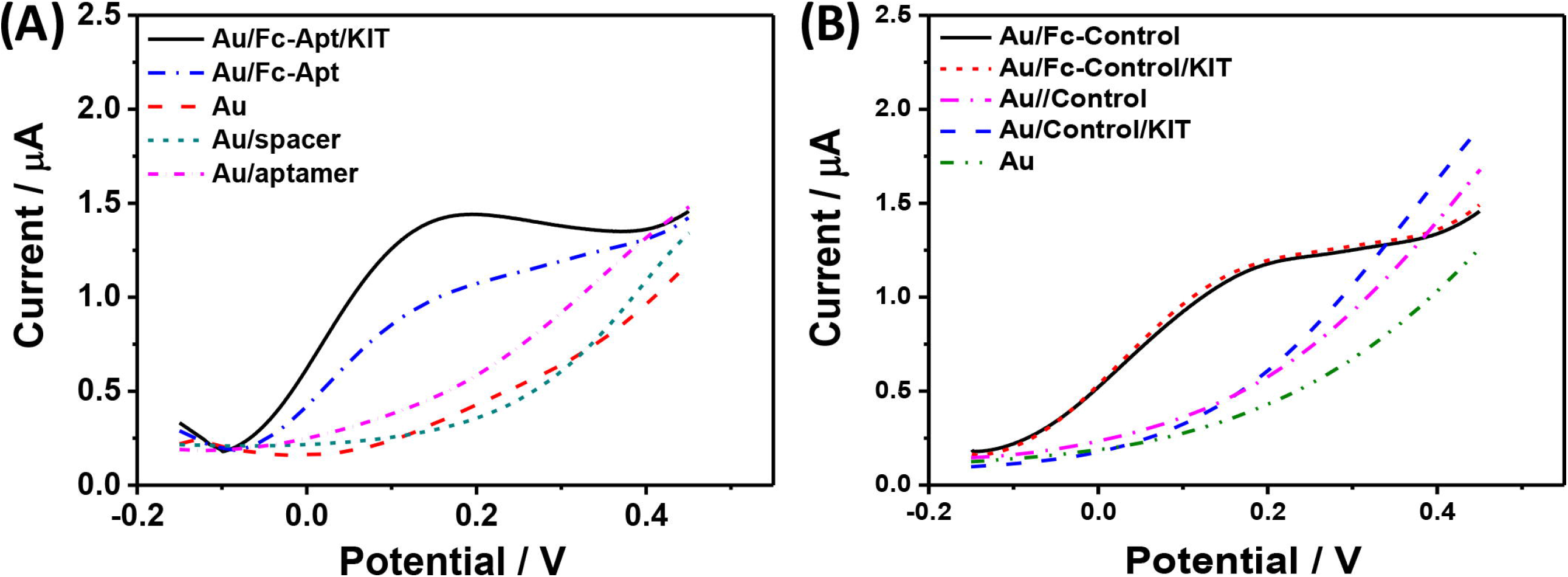
Stepwise atpasensor assembly with Fc reporter. **A**. Voltammograms of KIT electrode. **B**. Voltammograms of control oligo.

### 3.4. KIT detection using a conformation changing aptasensor

To assess the assay performance, KIT was incubated on the sensor with concentrations ranging from 10 pg/mL to 100 ng/mL and subsequently measured using SWV in PBS (pH = 7.4). In the presence of KIT, the Fc oxidation peak increased compared to the blank solution due to the direct electron-transfer of Fc resulting from the aptamer conformation change. As shown in **Fig. 4A**, the current response increased proportional to the KIT concentration. These data were compiled into a calibration curve (**Fig. 4B**) showing the linear relationship. The assay has a calculated limit of detection (LOD) of 1.15 pg/mL and a limit of quantification (LOQ) of 10.0 pg/mL with a linear dynamic range from 10 pg/mL to 100 ng/mL. The sensor reproducibility was evaluated by measuring 0.5 ng/mL of KIT with five independent electrodes. The coefficient of variation was 2.35%, indicating that the proposed sensor exhibited high reproducibility (**Supplemental Fig. 3**).

**Figure 4.**
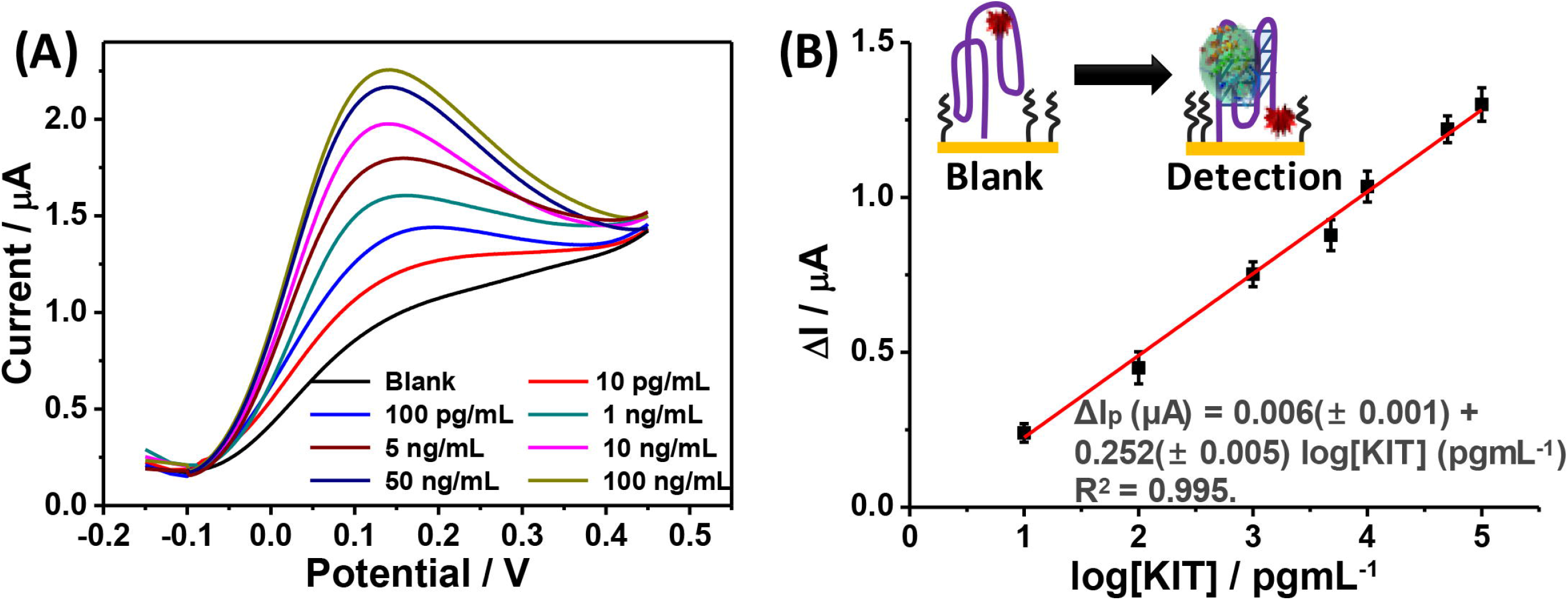
KIT conformation changing aptasensor. **A**. Voltammograms at various concentrations of KIT (10 pg/mL to 100 ng/mL) in 0.1 M PBS (pH 7.4). **B**. Calibration curve. All measurements were performed in triplicate where error bars indicate ±1σ.

### 3.5. Selectivity

Selectivity is an important characteristic of any receptor-target-based assay. While the specificity of the KIT aptamer was previously reported (Zhao et al., 2015; Banerjee et al., 2020), it is crucial to verify the specificity with the addition of the Fc reporter and the change to electrochemical rather than optical readout. To investigate the selectivity of the conformation changing aptasensor, voltammograms with various proteins (IL-6, UEA, BSA, Fibrinogen, and HSA) were measured, as shown in **Fig. 5A**. In all cases, the current responses of the interfering protein were significantly lower than 0.1 µg/mL of KIT, despite being present at an order of magnitude higher concentration. This indicates that the proposed sensor has good selectivity against off-target proteins. Next, the aptasensor was examined in human serum spiked with KIT towards an *in vivo* application. The serum was diluted 10-fold with PBS to mimic interstitial fluid and KIT was spiked in at concentrations of 10 pg/mL, 500 pg/mL, and 100 ng/mL. **Fig. 5B** shows the measured voltammograms where the current response gradually increased linearly proportional with the KIT concentration. The corresponding calibration curve is shown in **Fig. 5C** confirming the linear relationship These data demonstrate that the developed KIT aptasensor has applicability in clinically relevant biofluids with high levels of background interfering species.

**Figure 5.**
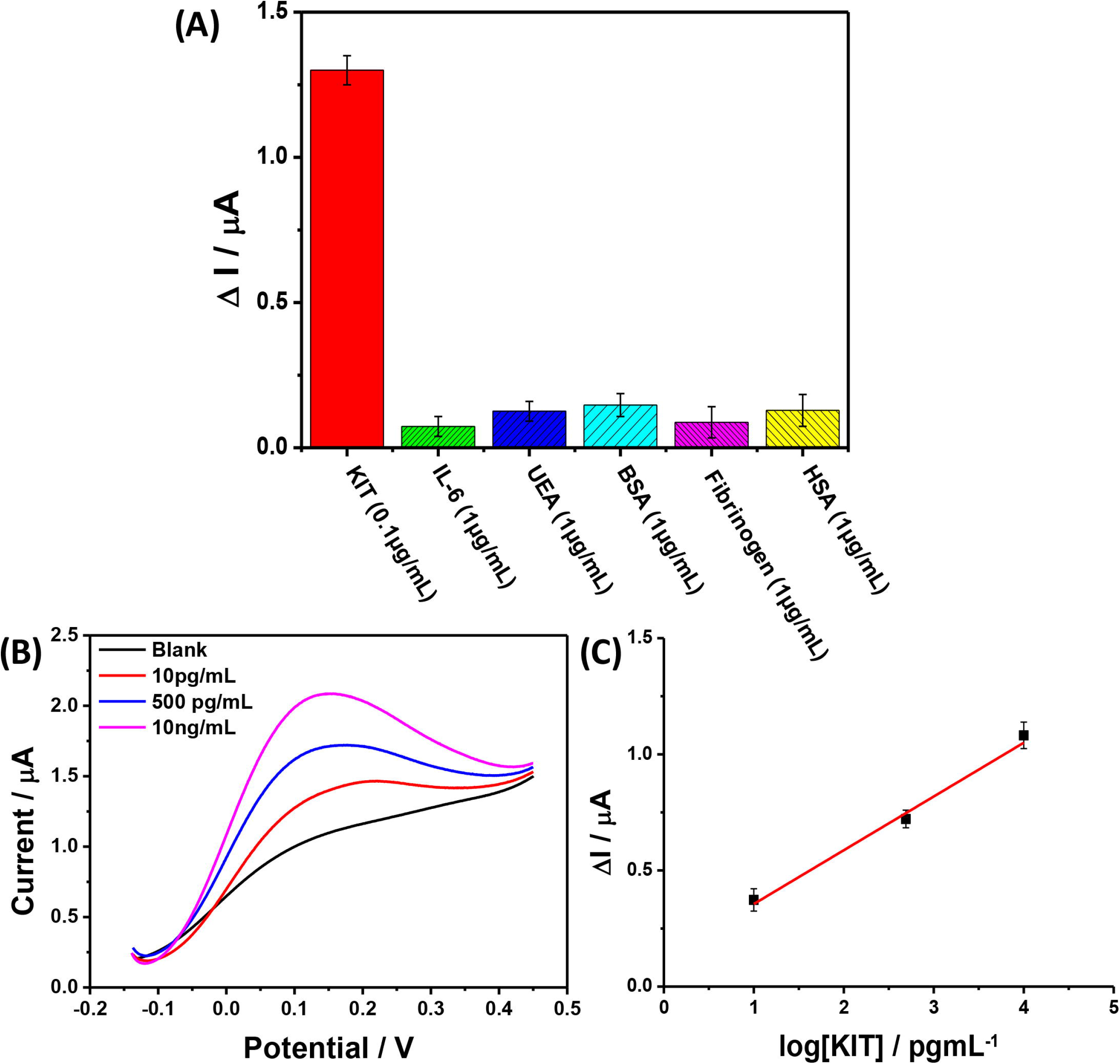
Selectivity of aptasensor. **A**. Comparison of aptasensor response to 0.1 ug/mL KIT and off-target proteins: IL-6, UEA, BSA, fibrinogen, and HAS at 1 ug/mL. **B**. Voltammograms recorded with spiked KIT in diluted human serum. **C**. Calibration curve. All measurements were performed in triplicate where error bars indicate ±1σ.

### 3.6. Cell culture media and serum assays

KIT is known to be overexpressed in GIST and Mastocytosis (Banerjee S et al. 2020). It has been shown that a soluble KIT fragment (sKIT) is released from cell surfaces and can be used for cancer theranostics (Akin et al., 2000; DePrimo et al., 2009; Sarlomo-Rikala et al., 1998). To investigate the clinical applicability of the reported KIT conformation changing aptasensor towards *in vivo* cancer treatment monitoring, we collected conditioned media containing sKIT from a GIST (GIST-T1) and a Systemic Mastocytosis (HMC-1.2) cell line, each of which has an oncogenic KIT mutation and overexpresses KIT protein (Ainsua-Enrich et al., 2015; Tabone et al., 2005). Conditioned media from a pancreatic cancer cell line, MIA-PaCa2, that does not express KIT was used as a control. Each supernatant was diluted 10^4^-fold in PBS solution and incubated on the aptasensors for 30 min (**Fig. 6A-B**). From the triplicate measurements, KIT concentrations were determined to be 12.84 ± 3.99 and 7.08 ± 1.98 ng/mL for HMC-1.2 and GIST-T1, respectively, whereas it was 0.06 ± 0.4 ng/mL for MIA-PaCa2 **(Fig. 6C**). These data are consistent with the western blot results run on the same samples (inset of **Fig. 6D**) (Banerjee et al., 2020). With electrochemical and western blot measurements, sKIT concentrations in the conditioned media of HMC-1.2 and GIST-T1 cells, which overexpress KIT, were significantly higher than MIA-PaCa2 cells. We also cross-validated our aptasensing results with a commercially available ELISA kit, where the three independent samples were analyzed. The results of the CM (background subtracted) for HMC-1.2, GIST-T1, and MIA-PaCa2 cells were 6.36 ± 0.28, 3.28 ± 0.55, and 0.03 ± 0.02 ng/mL, respectively. The sKIT levels detected using the aptasensor were higher than that of the ELISA possibly due to the higher affinity reagent and higher sensitivity of the electrochemical sensing approaches (Piro et al., 2016; Wang et al., 2017). To investigate the recovery of the KIT aptasensor, known concentrations of KIT (100 pg/mL, 1 ng/mL, and 10 ng/mL) were spiked in 10× diluted human serum. The percent recovery of spiked samples ranged from 85.9% to 117.5% (**Supplemental Table 1**). These measurements demonstrate that the developed conformation changing aptasensor has high selectivity and sensitivity to KIT.

**Figure 6.**
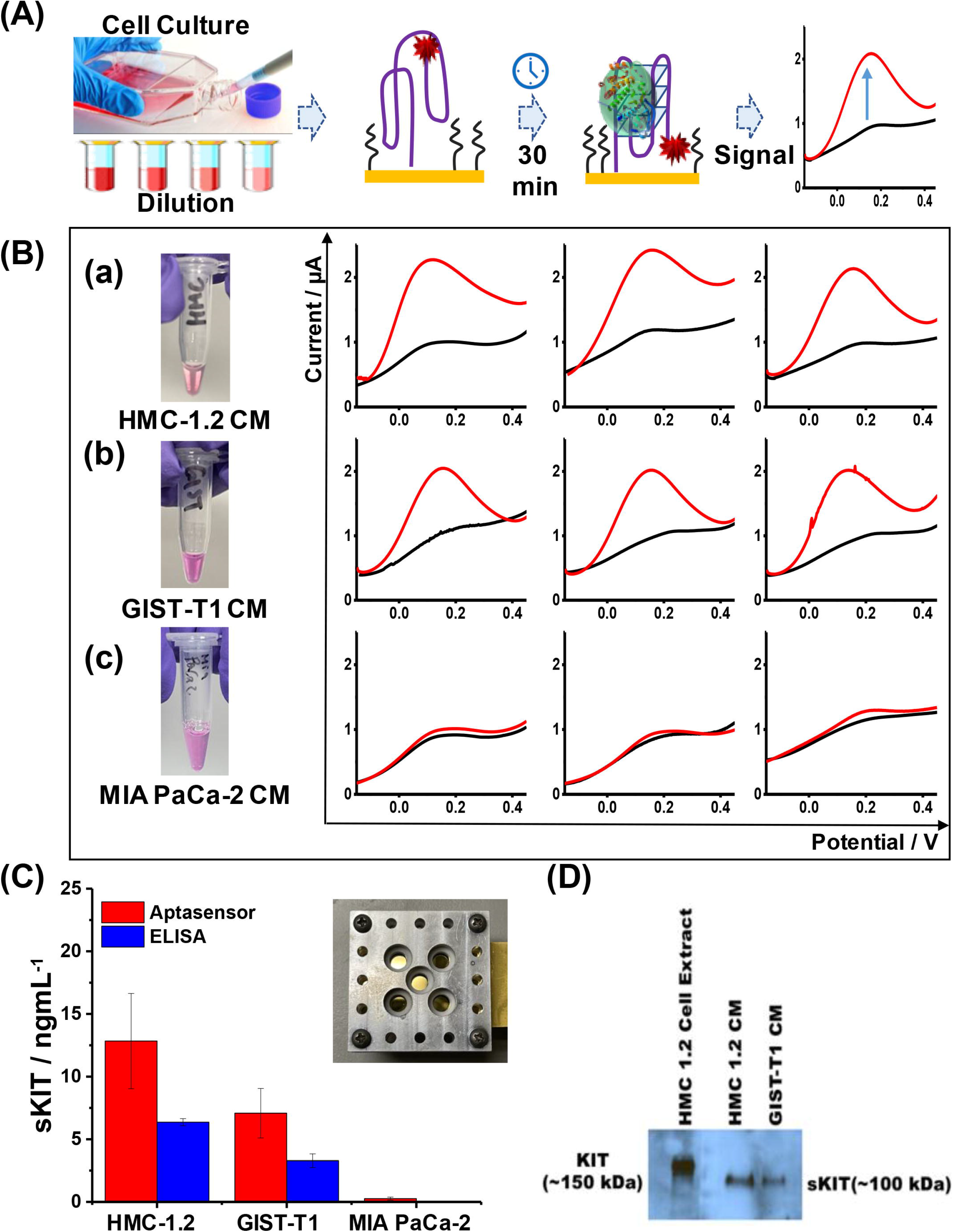
Clinical applicability. **(A)** Schematic and **(B)** response of the KIT aptasensor 30 min after incubation using conditioned medium of **(a)** HMC-1.2, **(b)** GIST-T1, and **(c)** MIA PaCa-2 cell lines. **(C)** Calculated sKIT concentration using aptasensor (red) and ELISA (blue) for each cell line. (Inset: Au electrode cells for KIT detection) **(D)** Western blot assay results of sKIT in the conditioned media.

## 4. Conclusion

In this work, we developed and characterized a conformation changing aptamer with a ferrocene reporter towards a GIST cancer biomarker, KIT. The stepwise sensor modification was investigated using CV and EIS, and experimental parameters such as the spacer ratio, aptamer immobilization time, pH, and KIT incubation time were optimized. In PBS, the aptasensor had a LOD of 1.15 pg/mL and a 10 pg/mL to 100 ng/mL linear dynamic range. The sensor was tested with off-target, interfering species and supernatant of conditioned medium with no appreciable non-specific binding. The proposed sensor was able to detect sKIT within 35 min unlike the traditional ELISA technique requiring several hours. The sensor successfully detected sKIT, which is released from the HMC-1.2 and GIST-T1 cells, which demonstrates that the reported aptasensor has both high specificity and high sensitivity. Therefore, the developed method can be applied as a fast routine laboratory test in CM, serum, and plasma samples obtained from clinical laboratories. Further, this work lays the foundation for a reagent-less assay towards *in vivo* treatment monitoring of KIT overexpressing cancers.

## Data Availability

Available upon request.

## Acknowledgments

This work was partially supported by the National Science Foundation (1621825), the National Institutes of Health (R41DA044905, R01 CA226803, and UL1TR001442), the UCSD Center for Wearable Systems (CWS), Samsung, the Food and Drug Administration (R01 FD006334), and GIST Research Fund. The content is solely the responsibility of the author(s) and does not necessarily represent the official views of the National Institutes of Health.

## References

Ainsua-Enrich, E., Serrano-Candelas, E., Álvarez-Errico, D., Picado, C., Sayós, J., Rivera, J., Martín, M., 2015. The Adaptor 3BP2 Is Required for KIT Receptor Expression and Human Mast Cell Survival. The Journal of Immunology 194, 4309–4318. https://doi.org/10.4049/jimmunol.1402887

Akin, C., Schwartz, L.B., Kitoh, T., Obayashi, H., Worobec, A.S., Scott, L.M., Metcalfe, D.D., 2000. Soluble stem cell factor receptor (CD117) and IL-2 receptor alpha chain (CD25) levels in the plasma of patients with mastocytosis: relationships to disease severity and bone marrow pathology. Blood 96, 1267–1273.

Aronoff-Spencer, E., Venkatesh, A.G., Sun, A., Brickner, H., Looney, D., Hall, D.A., 2016. Detection of Hepatitis C core antibody by dual-affinity yeast chimera and smartphone-based electrochemical sensing. Biosensors and Bioelectronics 86, 690–696. https://doi.org/10.1016/j.bios.2016.07.023

Arrieta, O., Cacho, B., Morales-Espinosa, D., Ruelas-Villavicencio, A., Flores-Estrada, D., Hernández-Pedro, N., 2007. The progressive elevation of alpha fetoprotein for the diagnosis of hepatocellular carcinoma in patients with liver cirrhosis. BMC Cancer 7, 28. https://doi.org/10.1186/1471-2407-7-28

Arya, S.K., Estrela, P., 2018. Recent Advances in Enhancement Strategies for Electrochemical ELISA-Based Immunoassays for Cancer Biomarker Detection. Sensors (Basel) 18. https://doi.org/10.3390/s18072010

Banerjee, S., Yoon, H., Yebra, M., Tang, C.-M., Gilardi, M., Shankara Narayanan, J.S., White, R.R., Sicklick, J.K., Ray, P., 2020. Anti-KIT DNA Aptamer for Targeted Labeling of Gastrointestinal Stromal Tumor. Mol. Cancer Ther. 19, 1173–1182. https://doi.org/10.1158/1535-7163.MCT-19-0959

Black, R.A., 2002. Tumor necrosis factor-α converting enzyme. The International Journal of Biochemistry & Cell Biology 34, 1–5. https://doi.org/10.1016/S1357-2725(01)00097-8

Bray, F., Ferlay, J., Soerjomataram, I., Siegel, R.L., Torre, L.A., Jemal, A., 2018. Global cancer statistics 2018: GLOBOCAN estimates of incidence and mortality worldwide for 36 cancers in 185 countries. CA: A Cancer Journal for Clinicians 68, 394–424. https://doi.org/10.3322/caac.21492

Cho, H., Yeh, E.-C., Sinha, R., Laurence, T.A., Bearinger, J.P., Lee, L.P., 2012. Single-Step Nanoplasmonic VEGF165 Aptasensor for Early Cancer Diagnosis. ACS Nano 6, 7607–7614. https://doi.org/10.1021/nn203833d

Chung, S., Moon, J.-M., Choi, J., Hwang, H., Shim, Y.-B., 2018. Magnetic force assisted electrochemical sensor for the detection of thrombin with aptamer-antibody sandwich formation. Biosens Bioelectron 117, 480–486. https://doi.org/10.1016/j.bios.2018.06.068

Collie, G.W., Parkinson, G.N., 2011. The application of DNA and RNA G-quadruplexes to therapeutic medicines. Chem. Soc. Rev. 40, 5867–5892. https://doi.org/10.1039/C1CS15067G

DePrimo, S.E., Huang, X., Blackstein, M.E., Garrett, C.R., Harmon, C.S., Schöffski, P., Shah, M.H., Verweij, J., Baum, C.M., Demetri, G.D., 2009. Circulating Levels of Soluble KIT Serve as a Biomarker for Clinical Outcome in Gastrointestinal Stromal Tumor Patients Receiving Sunitinib following Imatinib Failure. Clin Cancer Res 15, 5869–5877. https://doi.org/10.1158/1078-0432.CCR-08-2480

Ellington, A.D., Szostak, J.W., 1990. In vitro selection of RNA molecules that bind specific ligands. Nature 346, 818–822. https://doi.org/10.1038/346818a0

Feagin, T.A., Maganzini, N., Soh, H.T., 2018. Strategies for Creating Structure-Switching Aptamers. ACS Sens. 3, 1611–1615. https://doi.org/10.1021/acssensors.8b00516

Goossens, N., Nakagawa, S., Sun, X., Hoshida, Y., 2015. Cancer biomarker discovery and validation. Transl Cancer Res 4, 256–269. https://doi.org/10.3978/j.issn.2218-676X.2015.06.04

Harpio, R., Einarsson, R., 2004. S100 proteins as cancer biomarkers with focus on S100B in malignant melanoma. Clin Biochem 37, 512–518. https://doi.org/10.1016/j.clinbiochem.2004.05.012

Hianik, T., Wang, J., 2009. Electrochemical Aptasensors – Recent Achievements and Perspectives. Electroanalysis 21, 1223–1235. https://doi.org/10.1002/elan.200904566

Hristova, V.A., Chan, D.W., 2019. Cancer biomarker discovery and translation: proteomics and beyond. Expert Rev Proteomics 16, 93–103. https://doi.org/10.1080/14789450.2019.1559062

Kawakita, M., Yonemura, Y., Miyake, H., Ohkubo, T., Asou, N., Hayakawa, K., Nakamura, M., Kitoh, T., Osawa, H., Takatsuki, K., 1995. Soluble c-kit molecule in serum from healthy individuals and patients with haemopoietic disorders. British Journal of Haematology 91, 23–29. https://doi.org/10.1111/j.1365-2141.1995.tb05239.x

Kypr, J., Kejnovská, I., Renciuk, D., Vorlícková, M., 2009. Circular dichroism and conformational polymorphism of DNA. Nucleic Acids Res 37, 1713–1725. https://doi.org/10.1093/nar/gkp026

Lai, R.Y., Plaxco, K.W., Heeger, A.J., 2007. Aptamer-Based Electrochemical Detection of Picomolar Platelet-Derived Growth Factor Directly in Blood Serum. Anal. Chem. 79, 229–233. https://doi.org/10.1021/ac061592s

Lubin, A.A., Plaxco, K.W., 2010. Folding-Based Electrochemical Biosensors: The Case for Responsive Nucleic Acid Architectures. Acc. Chem. Res. 43, 496–505. https://doi.org/10.1021/ar900165x

Makowska, J.S., Cieslak, M., Kowalski, M.L., 2009. Stem cell factor and its soluble receptor (c-kit) in serum of asthmatic patients- correlation with disease severity. BMC Pulmonary Medicine 9, 27. https://doi.org/10.1186/1471-2466-9-27

Meirinho, S.G., Dias, L.G., Peres, A.M., Rodrigues, L.R., 2016. Voltammetric aptasensors for protein disease biomarkers detection: A review. Biotechnology Advances 34, 941–953. https://doi.org/10.1016/j.biotechadv.2016.05.006

Miettinen, M., Lasota, J., 2005. KIT (CD117): A Review on Expression in Normal and Neoplastic Tissues, and Mutations and Their Clinicopathologic Correlation. Applied Immunohistochemistry & Molecular Morphology 13, 205–220. https://doi.org/10.1097/01.pai.0000173054.83414.22

Moon, J.-M., Kim, D.-M., Kim, M.H., Han, J.-Y., Jung, D.-K., Shim, Y.-B., 2017. A disposable amperometric dual-sensor for the detection of hemoglobin and glycated hemoglobin in a finger prick blood sample. Biosens Bioelectron 91, 128–135. https://doi.org/10.1016/j.bios.2016.12.038

Naeem, M., Dahiya, M., Clark, J.I., Creech, S.D., Alkan, S., 2002. Analysis of c-kit protein expression in small-cell lung carcinoma and its implication for prognosis. Hum. Pathol. 33, 1182–1187. https://doi.org/10.1053/hupa.2002.129199

Nicholson, B.D., Shinkins, B., Pathiraja, I., Roberts, N.W., James, T.J., Mallett, S., Perera, R., Primrose, J.N., Mant, D., 2015. Blood CEA levels for detecting recurrent colorectal cancer. Cochrane Database of Systematic Reviews. https://doi.org/10.1002/14651858.CD011134.pub2

Oh, S.S., Plakos, K., Lou, X., Xiao, Y., Soh, H.T., 2010. In vitro selection of structure-switching, self-reporting aptamers. PNAS 107, 14053–14058. https://doi.org/10.1073/pnas.1009172107

Pilloni, L., Bianco, P., Difelice, E., Cabras, S., Castellanos, M.E., Atzori, L., Ferreli, C., Mulas, P., Nemolato, S., Faa, G., 2011. The usefulness of c-Kit in the immunohistochemical assessment of melanocytic lesions. Eur J Histochem 55. https://doi.org/10.4081/ejh.2011.e20

Piro, B., Shi, S., Reisberg, S., Noël, V., Anquetin, G., 2016. Comparison of Electrochemical Immunosensors and Aptasensors for Detection of Small Organic Molecules in Environment, Food Safety, Clinical and Public Security. Biosensors (Basel) 6. https://doi.org/10.3390/bios6010007

Plaxco, K.W., Soh, H.T., 2011. Switch-based biosensors: a new approach towards real-time, in vivo molecular detection. Trends in Biotechnology 29, 1–5. https://doi.org/10.1016/j.tibtech.2010.10.005

Poruk, K.E., Gay, D.Z., Brown, K., Mulvihill, J.D., Boucher, K.M., Scaife, C.L., Firpo, M.A., Mulvihill, S.J., 2013. The Clinical Utility of CA 19-9 in Pancreatic Adenocarcinoma: Diagnostic and Prognostic Updates. Curr Mol Med 13, 340–351.

Radi, A.-E., O’Sullivan, C.K., 2006. Aptamer conformational switch as sensitive electrochemical biosensor for potassium ion recognition. Chem. Commun. 3432–3434. https://doi.org/10.1039/B606804A

Ray, P., Viles, K.D., Soule, E.E., Woodruff, R.S., 2013. Application of Aptamers for Targeted Therapeutics. Arch. Immunol. Ther. Exp. 61, 255–271. https://doi.org/10.1007/s00005-013-0227-0

Reverri, E.J., Devitt, A.A., Kajzer, J.A., Baggs, G.E., Borschel, M.W., 2018. Review of the Clinical Experiences of Feeding Infants Formula Containing the Human Milk Oligosaccharide 2′- Fucosyllactose. Nutrients 10. https://doi.org/10.3390/nu10101346

Sarlomo-Rikala, M., Kovatich, A.J., Barusevicius, A., Miettinen, M., 1998. CD117: a sensitive marker for gastrointestinal stromal tumors that is more specific than CD34. Mod Pathol 11, 728–734.

Scholler, N., Urban, N., 2007. CA125 in Ovarian Cancer. Biomark Med 1, 513–523. https://doi.org/10.2217/17520363.1.4.513

Song, S., Wang, L., Li, J., Fan, C., Zhao, J., 2008. Aptamer-based biosensors. TrAC Trends in Analytical Chemistry 27, 108–117. https://doi.org/10.1016/j.trac.2007.12.004

Sun, A.C., Hall, D.A., 2019. Point-of-Care Smartphone-based Electrochemical Biosensing. Electroanalysis 31, 2–16. https://doi.org/10.1002/elan.201800474

Tabone, S., Théou, N., Wozniak, A., Saffroy, R., Deville, L., Julié, C., Callard, P., Lavergne-Slove, A., Debiec-Rychter, M., Lemoine, A., Emile, J.-F., 2005. KIT overexpression and amplification in gastrointestinal stromal tumors (GISTs). Biochim Biophys Acta 1741, 165–172. https://doi.org/10.1016/j.bbadis.2005.03.011

Taguchi, T., Sonobe, H., Toyonaga, S., Yamasaki, I., Shuin, T., Takano, A., Araki, K., Akimaru, K., Yuri, K., 2002. Conventional and Molecular Cytogenetic Characterization of a New Human Cell Line, GIST- T1, Established from Gastrointestinal Stromal Tumor. Laboratory Investigation 82, 663–665. https://doi.org/10.1038/labinvest.3780461

Tajima, F., Kawatani, T., Ishiga, K., Nanba, E., Kawasaki, H., 1998. Serum soluble c-kit receptor and expression of c-kit protein and mRNA in acute myeloid leukemia. European Journal of Haematology 60, 289–296. https://doi.org/10.1111/j.1600-0609.1998.tb01042.x

Tong, P., Zhang, L., Xu, J.-J., Chen, H.-Y., 2011. Simply amplified electrochemical aptasensor of Ochratoxin A based on exonuclease-catalyzed target recycling. Biosensors and Bioelectronics 29, 97–101. https://doi.org/10.1016/j.bios.2011.07.075

Tucker, W.O., Shum, K.T., Tanner, J.A., 2012. G-quadruplex DNA aptamers and their ligands: structure, function and application. Curr Pharm Des 18, 2014–2026. https://doi.org/10.2174/138161212799958477

Tuerk, C., Gold, L., 1990. Systematic evolution of ligands by exponential enrichment: RNA ligands to bacteriophage T4 DNA polymerase. Science 249, 505–510. https://doi.org/10.1126/science.2200121

Wang, S.X., Acha, D., Shah, A.J., Hills, F., Roitt, I., Demosthenous, A., Bayford, R.H., 2017. Detection of the tau protein in human serum by a sensitive four-electrode electrochemical biosensor. Biosensors and Bioelectronics 92, 482–488. https://doi.org/10.1016/j.bios.2016.10.077

Willner, I., Zayats, M., 2007. Electronic Aptamer-Based Sensors. Angewandte Chemie International Edition 46, 6408–6418. https://doi.org/10.1002/anie.200604524

Wypych, J., Bennett, L.G., Schwartz, M.G., Clogston, C.L., Lu, H.S., Broudy, V.C., Bartley, T.D., Parker, V.P., Langley, K.E., 1995. Soluble kit receptor in human serum. Blood 85, 66–73.

Xiao, Y., Lubin, A.A., Heeger, A.J., Plaxco, K.W., 2005. Label-Free Electronic Detection of Thrombin in Blood Serum by Using an Aptamer-Based Sensor. Angewandte Chemie 117, 5592–5595. https://doi.org/10.1002/ange.200500989

Zhao, N., Pei, S.-N., Qi, J., Zeng, Z., Iyer, S.P., Pei, L., Tung, C.-H., Zu, Y., 2015. Oligonucleotide aptamer-drug conjugates for targeted therapy of acute myeloid leukemia. Biomaterials 67, 42–51. https://doi.org/10.1016/j.biomaterials.2015.07.025

Zhong, H.-L., Lu, X.-Z., Chen, X.-M., Yang, X.-H., Zhang, H.-F., Zhou, undefined L., Wang, L., Cao, K.-J., Huang, J., 2010. Relationship between stem cell factor/c-kit expression in peripheral blood and blood pressure. Journal of Human Hypertension 24, 220–225. https://doi.org/10.1038/jhh.2009.62

