## Supplemental for "Development of a Soluble KIT (sKIT) Electrochemical Aptasensor For Cancer Theranostics"

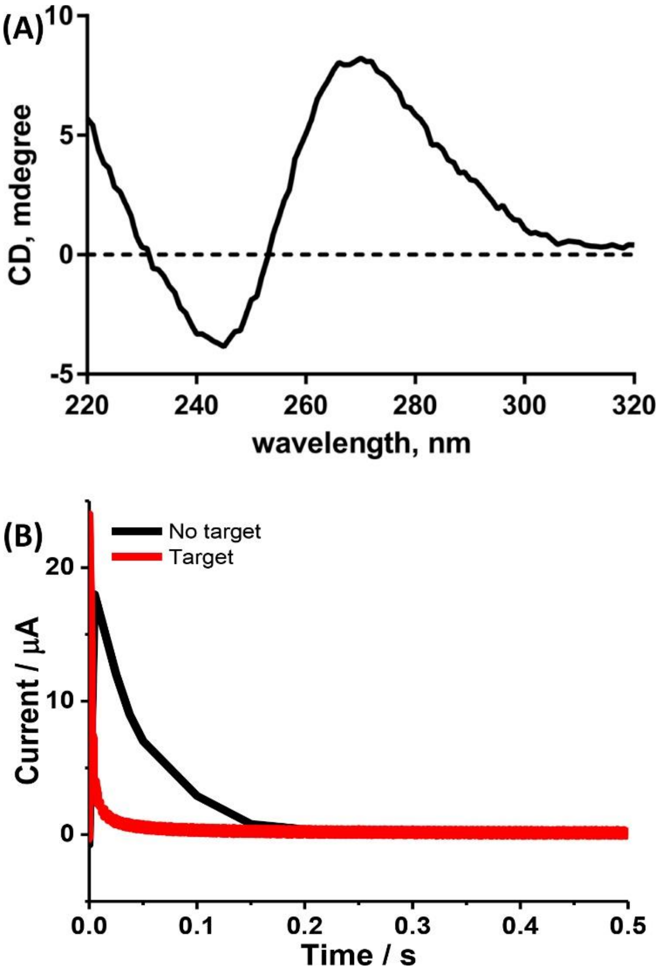


**Figure S1. A.** Circular dichroism (CD) spectroscopy of KIT aptamer **B.** Chronoamperometry recorded to confirm the difference in electron transfer rate between KIT bound and unbound aptamer where the aptamer has been modified at one of its ends with redox tag. Target binding increases electron transfer rate of redox tag, which translates into shorter decay times.

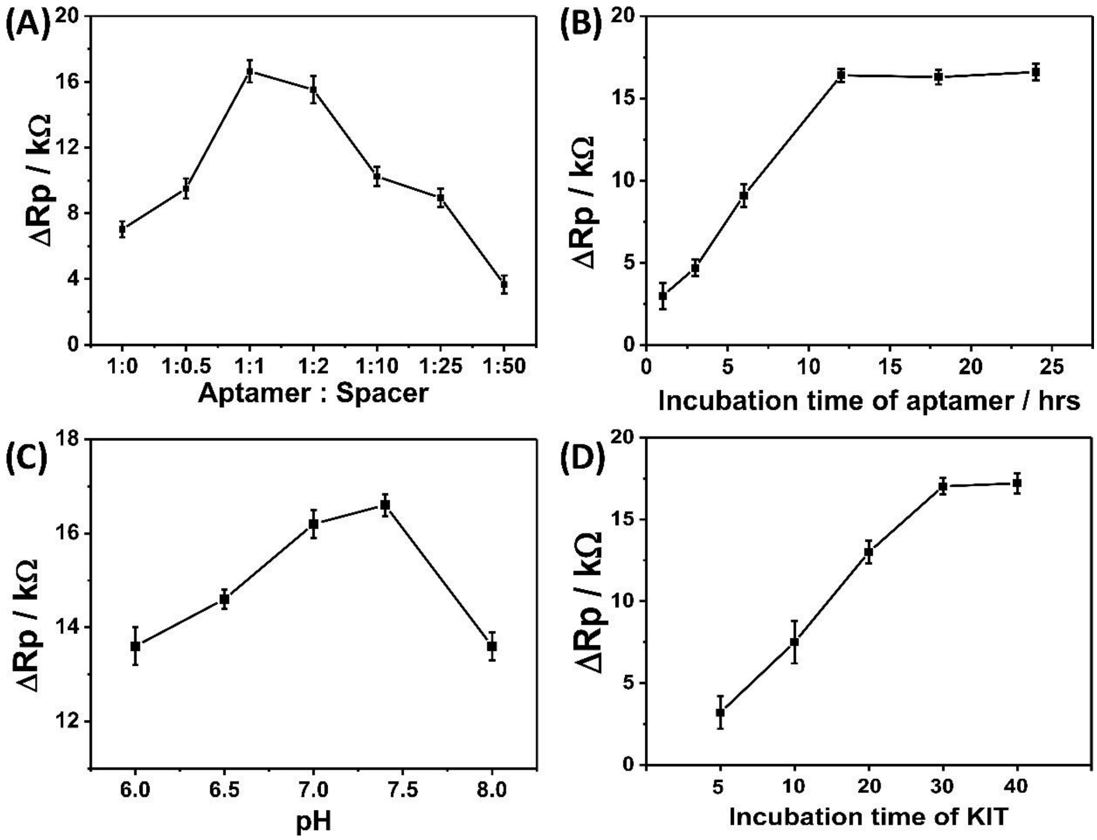


**Figure S2.** Optimization of experimental parameters in terms of the **A.** ratio of aptamer vs. spacer, **B.** incubation time of aptamer, **C.** pH, and **D.** incubation time of KIT using 10 ng/mL of KIT in 5 mM $Fe(CN)_6^{3-/4-}$ containing buffer.

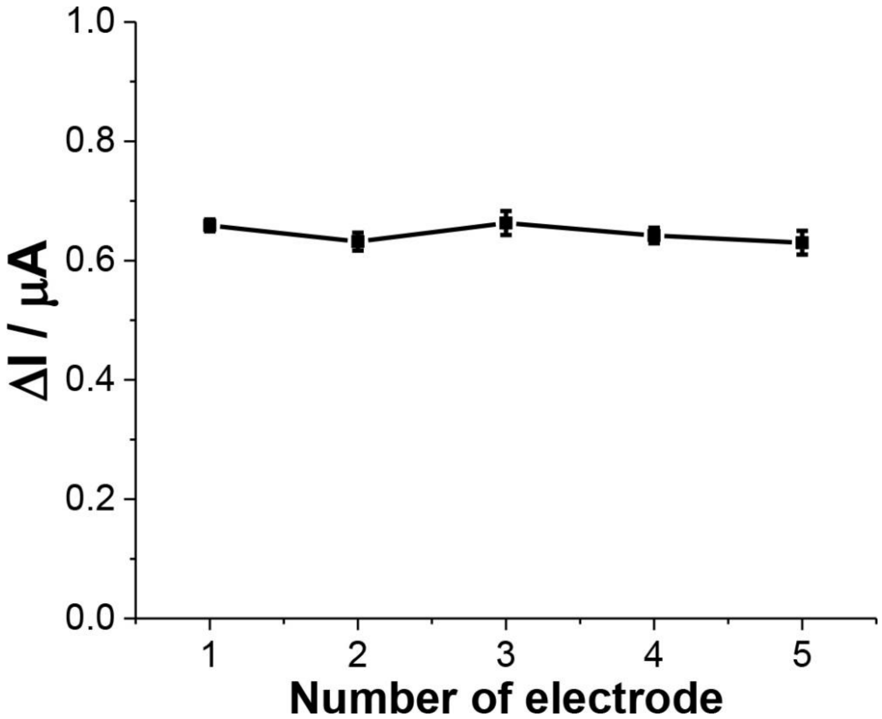


**Figure S3.** Reproducibility of proposed aptasensor (*n* = 5) toward KIT at 0.1 M PBS (pH 7.4).

**Table S1.** Recovery results of KIT in serum samples.

| Spiked (ng/mL) | Measured (ng/mL) | | | Recovery (%) | | |
|---|---|---|---|---|---|---|
| | Sample 1 | Sample 2 | Sample 3 | Sample 1 | Sample 2 | Sample 3 |
| 0.1 | 0.114 | 0.109 | 0.093 | 114.7 | 109.4 | 92.8 |
| 1 | 0.87 | 1.13 | 0.91 | 86.5 | 113.2 | 91.1 |
| 10 | 10.8 | 8.6 | 11.8 | 108.3 | 85.9 | 117.5 |
